# Measuring lactulose and mannitol levels using liquid chromatography coupled with tandem mass spectrum: application to clinical study of intestinal epithelium barrier function

**DOI:** 10.1101/2022.10.11.22280641

**Authors:** Lyvia M. V. C. Magalhães, Francisco A. P. Rodrigues, José Quirino-Filho, Rafhaella N.D.G. Gondim, Samilly Ribeiro, José K. Sousa, Marco Clementino, Bruna L.L. Maciel, Alexandre Havt, Armênio A. Santos, Pedro J.C. Magalhães, Aldo A.M. Lima

**Affiliations:** Center of Biomedicine and Department of Physiology and Pharmacology, Faculty of Medicine, Federal University of Ceará, Fortaleza, CE, Brazil; Department of Physical Education and Sport, Federal Institute of Education, Science and Technology of Ceará, Fortaleza, CE, Brazil; Nutrition Post-Graduation Program, Department of Nutrition, Federal University of Rio Grande do Norte, Natal, Rio Grande do Norte, Brazil

**Keywords:** Liquid chromatography coupled with tandem mass spectrum, lactulose:mannitol test validation, intestinal epithelium barrier function, children malnutrition

## Abstract

Lactulose and mannitol have been used to assess intestinal permeability and several methodologies have been used.

**Objectives:** This study aimed to validate the high-performance liquid chromatography method coupled with tandem mass spectrometry to measure mannitol and lactulose sugars.

**Material and Methods:** We used a high-performance liquid chromatography (HPLC) system coupled to an ABsciex Q-TRAP 5500 triple quadrupole mass spectrometer (MS/MS) with an ABSciex Electro Nebulization Interface (ESI) (Framingham, MA, USA). For the separation of lactulose and mannitol compounds in the HLPC, the analytical column HILIC-ZIC® from ES Industries (West Berlin, USA) was used. The parameters analyzed for analytical validation were specificity/selectivity, linearity, LD, LQ accuracy, precision (repeatability and intermediate precision) and matrix effect.

**Results:** The accuracy was demonstrated from the recovery at three concentration levels (100, 500 and 1000 ng/mL) and in triplicate, which showed recovery values above the recommended (>120%). Intermediate precision was determined at 24-hour intervals and the coefficients of variation found were less than 8.7%. The matrix effect was measured through the retention times in the standard samples and in the samples of the spiked standards in dilutions with urine samples, which varied between 99.3% and 100.3%. Urine samples from malnourished and healthy children were analyzed. The L:M ratio was considerably lower in the control group compared to the MN group (p<0.0001) and the mannitol excretion rate was higher (p<0.0001).

**Conclusions:** The results showed that the HPLC-MS/MS method was sensitive, specific, and accurate for the determination of molecular biomarkers of lactulose and mannitol. In addition, the L:M test is a functional test capable of determining with high sensitivity the barrier function damage of the intestinal epithelium in children with malnutrition compared to health control children.

## 1. Introduction

The need to show the quality of chemical measurements, through their comparability, traceability, and reliability, is being increasingly recognized and demanded. Unreliable analytical data can lead to disastrous decisions and irreparable financial losses. To ensure that a new analytical method generates reliable and interpretable information about the sample, it must undergo an evaluation called validation [1]. The lactulose:mannitol test (LM test) is a quantitative assay that measures two sugar molecules permeating across the functional intestinal epithelium barrier [2]. It consists of the oral administration of a solution containing lactulose and mannitol, followed by the collection of urine for a period (5 hours). Both sugars are absorbed through permeation through the functional intestinal epithelium barrier, are not metabolized in the body, and are excreted in the urine through the process of glomerular filtration. The monosaccharide mannitol (mw: 182 Da) due to its low molecular weight and hydrophilic characteristics, passes through the gastrointestinal functional barrier through hydrophilic pores via transcellular route. The disaccharide lactulose (mw: 342 Da) has a higher molecular weight and passes into the normal intestine in low amounts via the paracellular route. The reduction in villus length with consequent reduction in the absorption area reduces the absorption of mannitol, as well as the permeation of lactulose. On the other hand, the increase in intercellular space permeation or damage to the intestinal functional epithelium barrier results in increased absorption of lactulose. Thus, the lactulose:mannitol ratio is associated with changes in the absorption area, permeability, and damage to the intestinal functional epithelium barrier.

In this work we present the development of a high-performance liquid chromatography method coupled with tandem mass spectrometry (LC-MS/MS) to measure mannitol and lactulose sugars. In addition, we evaluate the functional intestinal epithelium barrier in children with malnutrition compared to health control children.

## 2. Material and methods

### 2.1 Standard and calibration solutions

Standard solutions of lactulose, mannitol, and sorbitol were prepared by dissolving them in eluent B (see below), containing acetonitrile and formic acid (0.05%) as the mobile phase. Individual stock solutions of each analyte were prepared at a concentration of 20 μg/mL and then were diluted in eluent B to 10, 50, 100, 500, 1000, 1500, and 2000 ng/mL. The final concentration of the internal standard (sorbitol) was 100 ng/mL **(Figure, Supplement Digital Content A)**.

### 2.2 HPLC-MS/MS for validation and measurement of lactulose and mannitol levels

The LC-MS/MS system consisted of a high-performance liquid chromatography instrument (Agilent, Santa Clara, CA, USA) with a 1200 series LC pump, degasser, autosampler, and column oven coupled to a Q-TRAP 5500 triple quadrupole mass spectrometer from ABsciex with an electrospray ionization interface (ESI) (ABSciex, Framingham, MA, USA). For the separation of the compounds by LC, a HILIC-ZIC® analytical column from (ES Industries, West Berlin, NJ, USA) was used. MS/MS parameters for monitored ion transitions were obtained using 10 ng/mL solutions of each substance at a flow rate of 10 μL/min in the negative ionization mode.

The mobile phase for chromatographic separation was composed of an aqueous fraction (eluent A) and an organic fraction (eluent B). The aqueous fraction contained a mixture of a 5.0 mmol/L ammonium acetate solution, and the organic fraction contained acetonitrile acidified with 0.05% formic acid. The mobile phase composition was altered for gradient elution based on the study by Kubica et al. [3] (**Table, Supplement Digital Content A**). The parameters analyzed for analytical validation were specificity, selectivity, linearity, limit of detection (LD), limit of quantification (LQ), accuracy, precision (repeatability and intermediate precision), and matrix effect.

### 2.3 Preparation of standard solutions and urine samples

Each 500 μL urine sample was initially diluted in 500 μL of ultrapure water, and 100 mg of Amberlite MB150 resin ion exchange was added to the diluted samples to eliminate excess sodium ions [6]. The samples were vortexed for 3 min and then centrifuged for 6 min at 9,000 rpm. Thereafter, 10 μL of the supernatant was added to 100 μL of the internal standard sorbitol solution (100 ng/mL) and diluted again in eluent B to a total volume of 2 mL. Lactulose, mannitol, and sorbitol were dissolved in four urine samples free of compounds of interest at a concentration of 20 μg/mL each. The enriched samples were prepared as previously described [3]. The final concentrations of the samples were 1000, 1500, 3000, and 6000 ng/mL for each substance. The prepared spiked samples were analyzed using LC-MS/MS.

### 2.4 Preparation and performance of the lactulose:mannitol test

After fasting for at least 5 h, the children ingested lactulose (5 g) and mannitol (1 g) diluted in 20 mL of water. Children weighing less than 10 kg ingested 2 mL/kg of this solution. We then determined the amount of lactulose and mannitol recovered from a urine sample taken in 5 h with 50 μL of 20 μg/mL chlorhexidine solution added per each 50 mL of urine as a preservative. Lactulose an mannitol analytes excreted in the urine samples were measured using LC-MS/MS.

### 2.5 Study design, population, and ethical approval

To validate in clinical specimens in the LC-MS/MS method, we used urine samples from intestinal permeability tests done in children. The clinical study has been performed on 34 children enrolled in the study protocol from the malnourished clinic IPREDE and Canarinho day care center at Fortaleza, Ceará, Brazil. Ethical approval of the study was provided by Federal University of Ceará and University of Virginia and informed consent and assent were provided by all participants. This case-control study was carried out in Fortaleza, CE, in the Northeast region of Brazil from August 19, 2010, to May 23, 2017. Malnourished (MN) children (weight-for-age Z-score [WAZ] < −2) were followed up at the Institute for the Promotion of Nutrition and Human Development for nutritional counseling. As a control group, healthy children from the Canarinho day care center were recruited in Fortaleza. All children were examined with permission from the mother or primary caregiver responsible for legal custody of the child. The study protocol and consent form were approved by the Institutional Review Board of the Federal University of Ceará and the National Council for Ethics in Research. We included children aged 0.5–7 years. Malnourished children were defined as those having a WAZ of less than −2 and healthy children (without any specific illness or fever) with a WAZ above −1 [4]. Healthy children were also defined as having a Z-score of WAZ > −1. Exclusion criteria defined in the study protocol were prolonged hospitalization or serious health issues, such as human immunodeficiency virus infection, tuberculosis, neonatal illness, kidney disease, or other illnesses diagnosed by a physician. In addition, children with a parent or a primary caregiver with cognitive deficits or younger than 16 years of age were excluded.

### 2.6 Sample size and statistical analysis

The lactulose:mannitol urinary excretion ratio was selected as the primary outcome variable and used to calculate the sample size for LM test validation. To detect a 30% change in the lactulose:mannitol ratio at a level of *P* = 0.05 and power 80%, we calculated that we would need 17 participants in each subgroup. This calculation was based on the data from preliminary studies in the same population (lactulose:mannitol ratio = 0.13 ± 0.04) [2]. Data were entered into spreadsheets (Microsoft Access software; Microsoft Corporation, Redmond, WA, USA) and verified by two independent researchers to ensure accuracy. All data were de-identified, and statistical analysis was performed using SPSS Statistics 20.0 (IBMCorporation, https://www.ibm.com). For analytical processing and measurement of mannitol and lactulose biomarker levels using LC-MS/MS, we used Analyst 1.4.1 software (http://www.appliedbiosystems.com). We used the Shapiro-Wilk test to assess the normality of the data distribution and Levene’s test to assess the equality of variances. For nonparametric variables, we used the Mann-Whitney U test for the independent samples. For qualitative variables, we used the χ2 test or Fisher’s exact test. We used analysis of covariance (ANCOVA) to explore the influence of children’s age on the differences in the lactulose:mannitol ratio between healthy children and children with MN **(Table, Supplement Digital Content E)**. The results were considered significant if *P* < 0.05.

## 3. Results

The identification of the precursor ions of the standard lactulose, mannitol, and sorbitol analytes using the automatic method of direct multiple reaction monitoring in the mass spectrum is shown in **Table A** and summarizes the results obtained from monitoring and analyzing MS/MS spectra of precursor and product ions of the injected standards of lactulose, mannitol, and sorbitol. The component ions of interest were detected in the negative ESI mode. The highest intensity product ions in the detector were 160,952 m/z, 112,798 m/z, and 112,912 for lactulose, mannitol, and sorbitol, respectively. The automatic parameters detected for precursor and product ions of the lactulose, mannitol, and sorbitol standards were determined by the flow injection analysis (FIA) coupled with LC (without the addition of the chromatographic column) and MS. The parameters selected for the FIA by the best signal strength of the product ions in the MS detector are listed in **Table, Supplement Digital Content B. Table B** shows the chromatographic conditions, when using the HILIC-ZIC^®^ column, and selected FIA and MS/MS parameters, as well as product ions best quantified and selected from the precursor ions of lactulose, mannitol, and sorbitol standards based on calibration curves and correlation coefficients.

**Table A.**
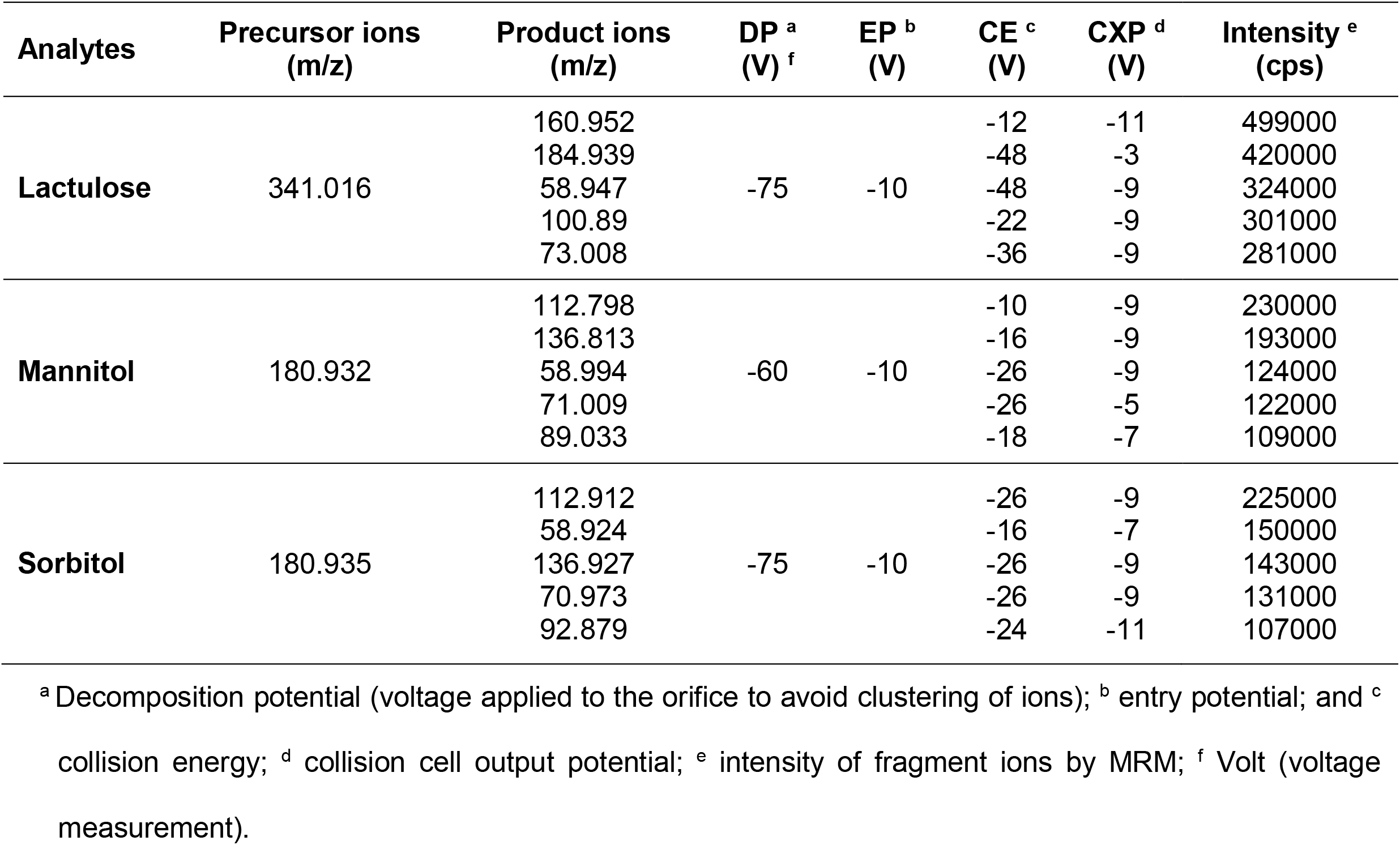
Mass spectrum monitoring and operational parameters of precursor ions and product ions of lactulose, mannitol, and sorbitol, through Multiple Reaction Monitoring (MRM).

**Table B.**
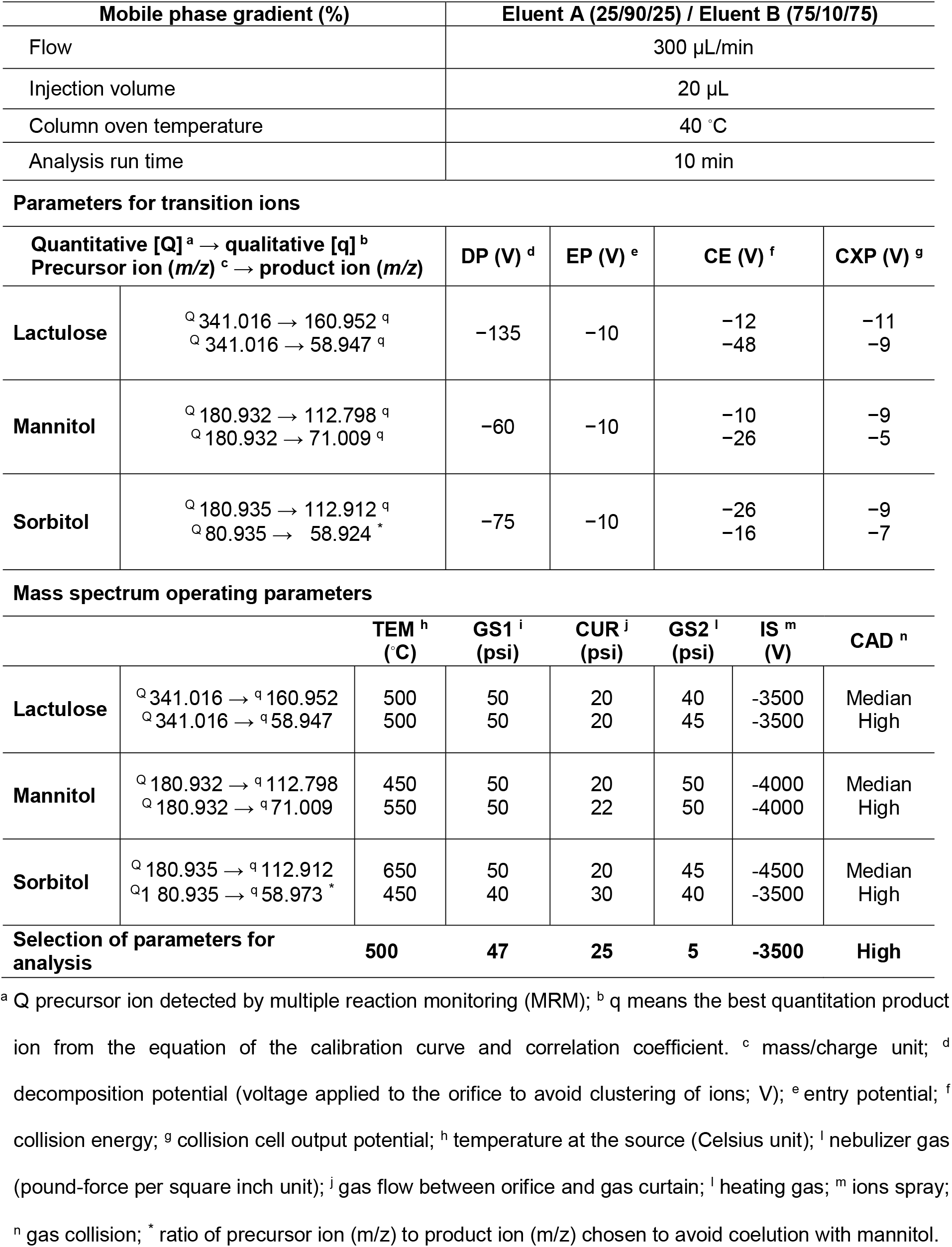
Column chromatographic conditions (HILIC-ZIC^®^) and optimal parameters for monitoring transition ions.

| Mobile phase gradient (%) | Eluent A (25/90/25) / Eluent B (75/10/75) |
| --- | --- |
| Flow | 300 µL/min |
| Injection volume | 20 µL |
| Column oven temperature | 40 °C |
| Analysis run time | 10 min |

**Parameters for transition ions**
| Quantitative [Q]<sup>a</sup> → qualitative [q]<sup>b</sup><br>Precursor ion (m/z)<sup>c</sup> → product ion (m/z) |  | DP (V)<sup>d</sup> | EP (V)<sup>e</sup> | CE (V)<sup>f</sup> | CXP (V)<sup>g</sup> |
| --- | --- | --- | --- | --- | --- |
| **Lactulose** | <sup>Q</sup> 341.016 → 160.952 <sup>q</sup><br><sup>Q</sup> 341.016 → 58.947 <sup>q</sup> | -135 | -10 | -12<br>-48 | -11<br>-9 |
| **Mannitol** | <sup>Q</sup> 180.932 → 112.798 <sup>q</sup><br><sup>Q</sup> 180.932 → 71.009 <sup>q</sup> | -60 | -10 | -10<br>-26 | -9<br>-5 |
| **Sorbitol** | <sup>Q</sup> 180.935 → 112.912 <sup>q</sup><br><sup>Q</sup> 80.935 → 58.924<sup>*</sup> | -75 | -10 | -26<br>-16 | -9<br>-7 |

**Mass spectrum operating parameters**
|  |  | TEM<sup>h</sup><br>(°C) | GS1<sup>i</sup><br>(psi) | CUR<sup>j</sup><br>(psi) | GS2<sup>l</sup><br>(psi) | IS<sup>m</sup><br>(V) | CAD<sup>n</sup> |
| --- | --- | --- | --- | --- | --- | --- | --- |
| **Lactulose** | <sup>Q</sup> 341.016 → <sup>q</sup> 160.952<br><sup>Q</sup> 341.016 → <sup>q</sup> 58.947 | 500<br>500 | 50<br>50 | 20<br>20 | 40<br>45 | -3500<br>-3500 | Median<br>High |
| **Mannitol** | <sup>Q</sup> 180.932 → <sup>q</sup> 112.798<br><sup>Q</sup> 180.932 → <sup>q</sup> 71.009 | 450<br>550 | 50<br>50 | 20<br>22 | 50<br>50 | -4000<br>-4000 | Median<br>High |
| **Sorbitol** | <sup>Q</sup> 180.935 → <sup>q</sup> 112.912<br><sup>Q</sup> 180.935 → <sup>q</sup> 58.973<sup>*</sup> | 650<br>450 | 50<br>40 | 20<br>30 | 45<br>40 | -4500<br>-3500 | Median<br>High |
| **Selection of parameters for analysis** |  | **500** | **47** | **25** | **5** | **-3500** | **High** |

<sup>a</sup> Q precursor ion detected by multiple reaction monitoring (MRM); <sup>b</sup> q means the best quantitation product ion from the equation of the calibration curve and correlation coefficient. <sup>c</sup> mass/charge unit; <sup>d</sup> decomposition potential (voltage applied to the orifice to avoid clustering of ions; V); <sup>e</sup> entry potential; <sup>f</sup> collision energy; <sup>g</sup> collision cell output potential; <sup>h</sup> temperature at the source (Celsius unit); <sup>i</sup> nebulizer gas (pound-force per square inch unit); <sup>j</sup> gas flow between orifice and gas curtain; <sup>l</sup> heating gas; <sup>m</sup> ions spray; <sup>n</sup> gas collision; <sup>\*</sup> ratio of precursor ion (m/z) to product ion (m/z) chosen to avoid coelution with mannitol.

**Table C** shows the precursor and product ions of the lactulose, mannitol, and sorbitol standard analytes with their respective calibration curve equations, detection and quantification limits, and correlation coefficients. The linear parts of the standard lactulose, mannitol, and sorbitol analyte curves were in the concentration range between 10 and 2000 ng/mL The correlation coefficients of the linear equations obtained for the three sugars were greater than 0.99. The calculations of LD and LQ were based on the standard deviation of the sample at a concentration of 100 ng/mL (the lowest concentration at which the method used was accurate and precise for each analyte) and the slope of the calibration curve in the region between 10 and 2000 ng/mL. The accuracy of the analytical method determined from the recovery and coefficients of variation of the standard analytes is summarized in detail in **Tables D and E, Table, Supplement Digital Content C**. The matrix effect interference of the standard analytes diluted in urine samples is shown in **Table, Supplement Digital Content D**.

**Table C.**
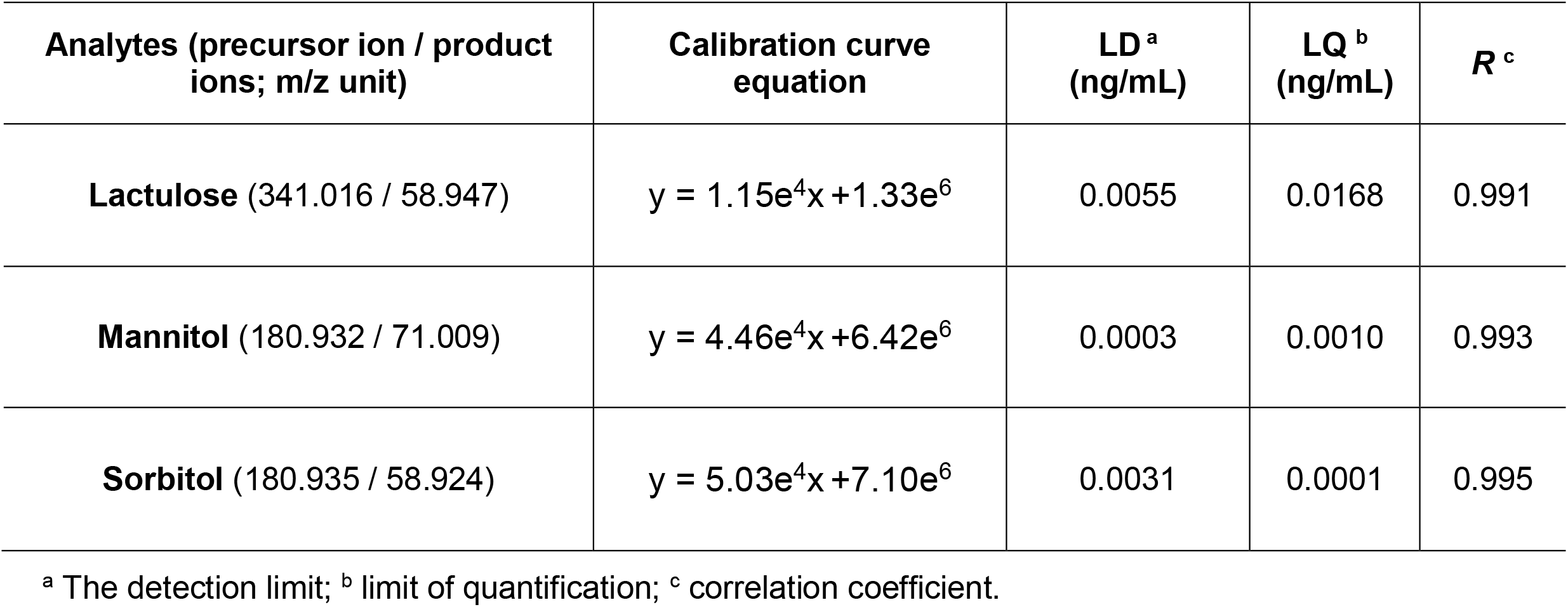
Linearity, limit of detection (LD), limit of quantification (LQ) of the method in the LC-MS/MS system for analysis of the excretion of lactulose, mannitol and sorbitol sugars.

**Table D.**
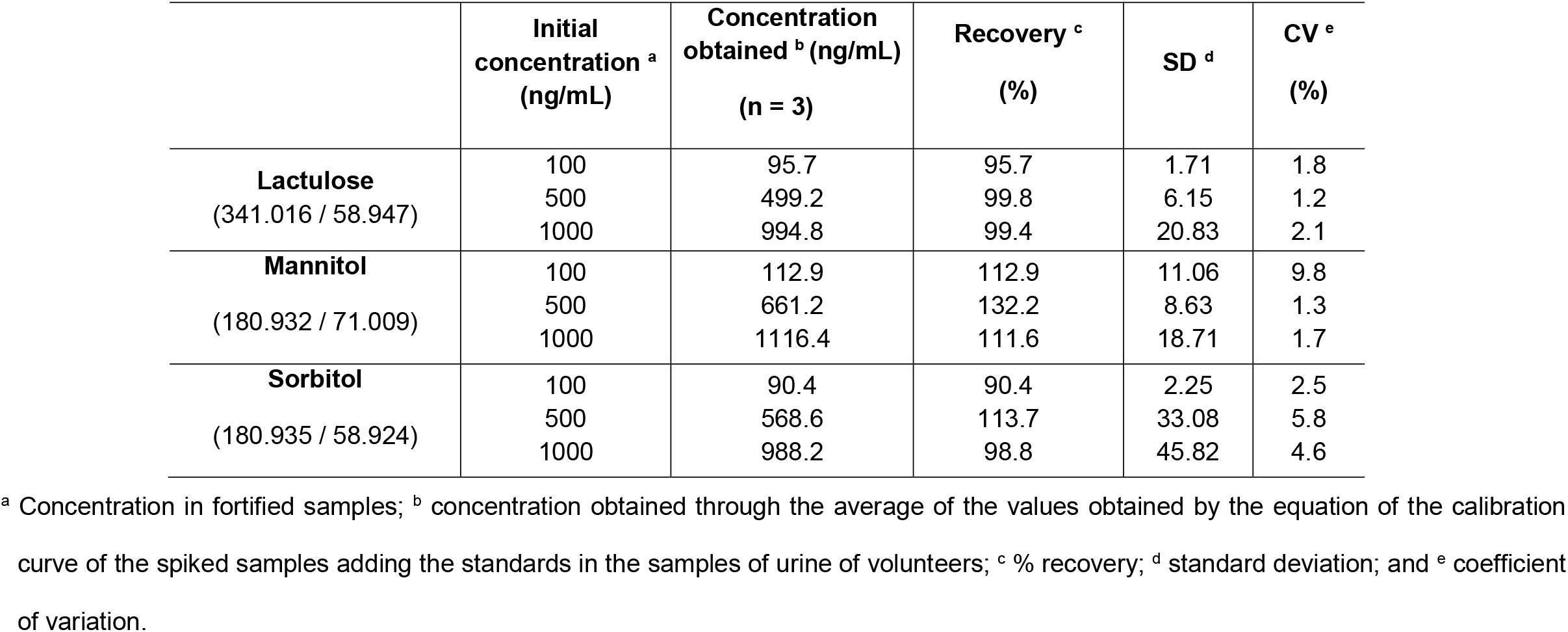
Repeatability of the method in the LC-MS/MS system to analyze the excretion of lactulose, mannitol, and sorbitol sugars.

**Table E.**
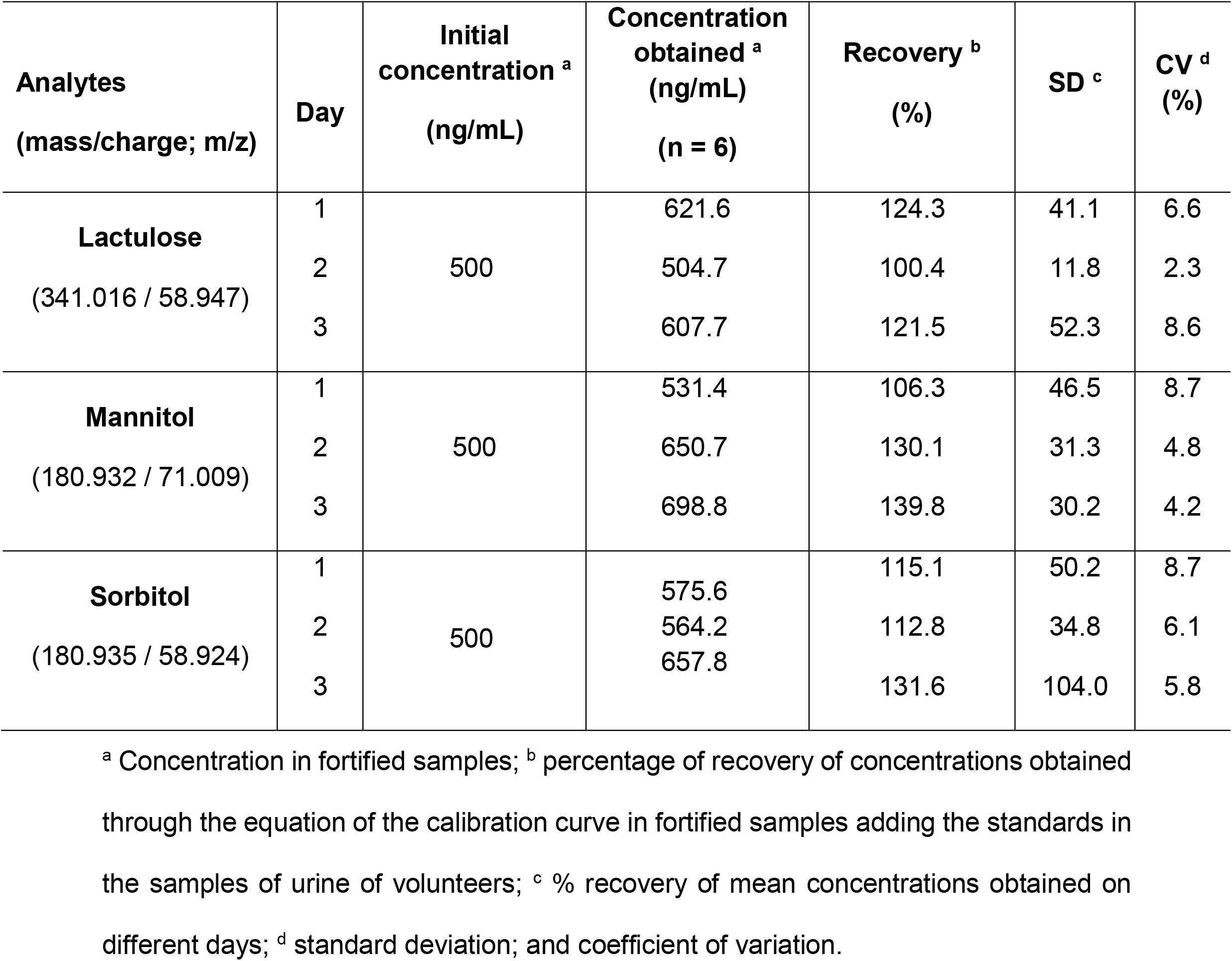
Intermediate precision of the method in the LC-MS/MS system for analysis of lactulose, mannitol, and sorbitol in urine samples.

The Mann whitney test was conducted to compare the differences between the two groups in the lactulose:mannitol ratio. The differences between the experimental groups in the L:M ratio and mannitol excretion is significant **(Figure A)**.

**Figure A.**
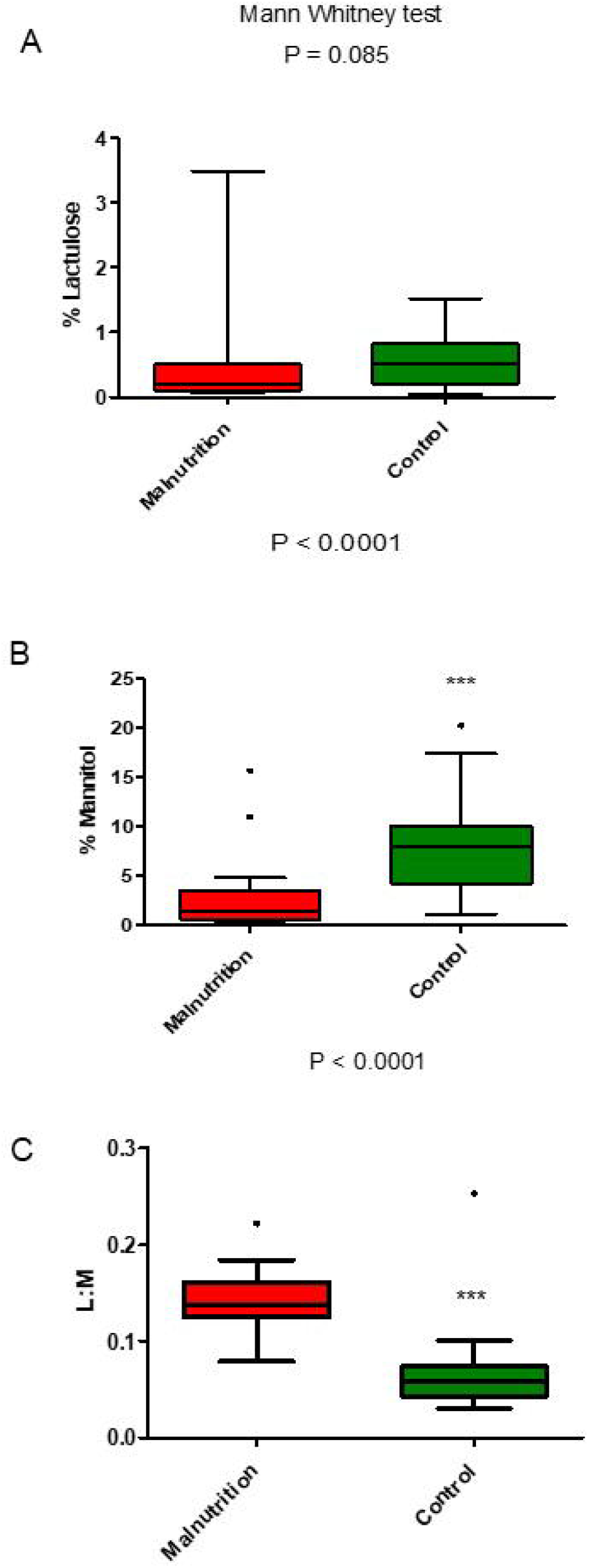
The distribution parameters associated with LM test in children are shown in the Boxplot plots. **Figure A.1** shows the Boxplot distribution of percentage of urinary lactulose excretion in the groups of healthy control and malnourished (MN) children. In **Figure A.2** we observe the parameter of the percentage of urinary excretion of mannitol in the groups of control and MN children. **Figure A.3** shows the lactulose:mannitol urinary excretion rate in the control and MN groups of children. The statistical test used for comparisons between the experimental groups and the P values when significant are shown in the graph. Details on understanding the figure are described in the manuscript text.

## 4. Discussion

Several chromatographic methods have been used for the analysis of lactulose and mannitol in urine [9,10]. However, the LC-MS/MS method proposed in this study has advantages over LC alone because of its higher sensitivity, specificity, and throughput (shorter sample run time) in the analysis of lactulose and mannitol as biomarkers [9].

The linearity of the method was demonstrated by the addition of known amounts of standards (lactulose, mannitol, and sorbitol). The calibration curves for the working ranges of the three analytes showed satisfactory linearity (R ≥ 0.991).

There are several approaches to assess accuracy and precision (repeatability and intermediate precision). Most commonly, the percentage of analyte recovery is calculated [10]. The coefficients of variation did not exceed 9.79%, below the limit accepted and suggested by Barboza et al. [2]. The recovery values obtained in order to assess the accuracy and repeatability of the method varied between 95.7 and 132.9, according to the concentrations analyzed. For intermediate precision, the recovery ranged between 100.94 and 139.76. Although recoveries close to 100% are desirable, the extent of analyte recovery up to 50–60% may be acceptable if the recovery is accurate and reproducible [10]. The accuracy and precision parameters are key to the validation process. They are required for all method validation studies, except for those with a qualitative purpose and intended only to demonstrate the presence of an analyte. For the quantitative analysis of trace elements, it is necessary to validate the detection and quantification limits. For the qualitative analysis, it is mandatory to validate only the detection limit. Linearity parameter evaluation is mandatory for new methods and qualitative analyses. The addition of a specific amount of ion-exchange resin to urine samples was sufficient to obtain high recovery values, as reported by Kubica et al. [3]. For further detail discussion in the LC-MS/MS analytical method please see the **Text, Supplemental Digital Content A**.

The lactulose:mannitol urinary excretion ratio test has recently been considered one of the best noninvasive tests to assess the area of absorption, permeability, and damage to the FGB [9,11]. In this study, we developed and validated a new robust, sensitive, specific, and accurate HPLC-MS/MS method for measuring sugar biomarkers, such as lactulose and mannitol. The lactulose:mannitol permeability test was considered abnormal or positive for comparison purposes if the LM urine excretion ratio was 0.0864 [14]. In particular, the rate of urinary mannitol excretion was significantly higher in the control group than in the MN group (*p*<0,0001). The rate of urinary lactulose excretion was ower in the groups of children MN than in the control group (*p* = 0.085). The L:M ratio was higher in the MN group than compared to the control group (*p*<0.001). Thus, the noninvasive test to assess the urinary excretion rates of lactulose and mannitol was consistent and therefore, may be applicable for the study of changes in the FGB as an ideal method for follow-up and evaluation of the response to preventive and curative interventions [15–16,18]. This study has advantages, such as the use of a robust, highly sensitive, specific, and accurate method for determining lactulose and mannitol levels. The limitation of the method is the total time of urine collection during 5 h, which makes its reproduction difficult. Our preliminary data show that the total collection time can be shortened to 1.5–2 h.

## 5. Conclusions

The results of this study allow us to propose LC-MS/MS as a robust, sensitive, specific, and accurate method for the determination of lactulose and mannitol to study the barrier function damage of the intestinal epithelium in children with malnutrition compared to health control children.

## Supporting information

Figure Supplement Digital Content A

Table, Supplement Digital Content A

Table, Supplement Digital Content B

Table, Supplement Digital Content C

Table, Supplement Digital Content D

Table, Supplement Digital Content E

Text, Supplement Digital Content A

## Data Availability

All data produced in the present work are contained in the manuscript

## Acknowledgement

The authors of this study are immensely grateful to the parents and guardians and participating children for signing the terms of free and arbitrary consent, acceptance, and cooperation in the study protocol.

The authors declare that they have no known competing commercial interests or personal relationships that could have appeared to influence the work reported in this paper.

## Author Contributions

Conceptualization, L.M.V.C. Magalhães, F. A. P. Rodrigues, R.N.D.G. Gondim, A.A. Santos, P.J.C. Magalhães, B.L.L. Maciel, A. Havt, A.A.M. Lima; methodology, F. A. P. Rodrigues, L.M.V.C. Magalhães, B.L.L. Maciel, R.N.D.G. Gondim, A. Havt, A.A.M. Lima; formal analysis, L.M.V.C. Magalhães, F. A. P. Rodrigues, A. Havt, A.A.M. Lima; data curation, L.M.V.C. Magalhães, F. A. P. Rodrigues, J. Quirino-Filho, A. Havt, A.A.M. Lima; writing-original draft preparation, L.M.V.C. Magalhães, F. A. P. Rodrigues, R.N.D.G. Gondim, S. Ribeiro, J. K. Sousa, M. Clementino, A.A. Santos, P.J.C. Magalhães, B.L.L. Maciel, A. Havt, A.A.M. Lima; writing-review and editing, L.M.V.C. Magalhães, F. A. P. Rodrigues, A.A. Santos, R.N.D.G. Gondim, S. Ribeiro, J. K. Sousa, M. Clementino, P.J.C. Magalhães, A. Havt, B.L.L. Maciel, A.A.M. Lima. All authors have read and agreed to the published version of the manuscript.

