## Supplementary figures and images for "Measuring lactulose and mannitol levels using liquid chromatography coupled with tandem mass spectrum: application to clinical study of intestinal epithelium barrier function"

### Figure Supplement Digital Content A

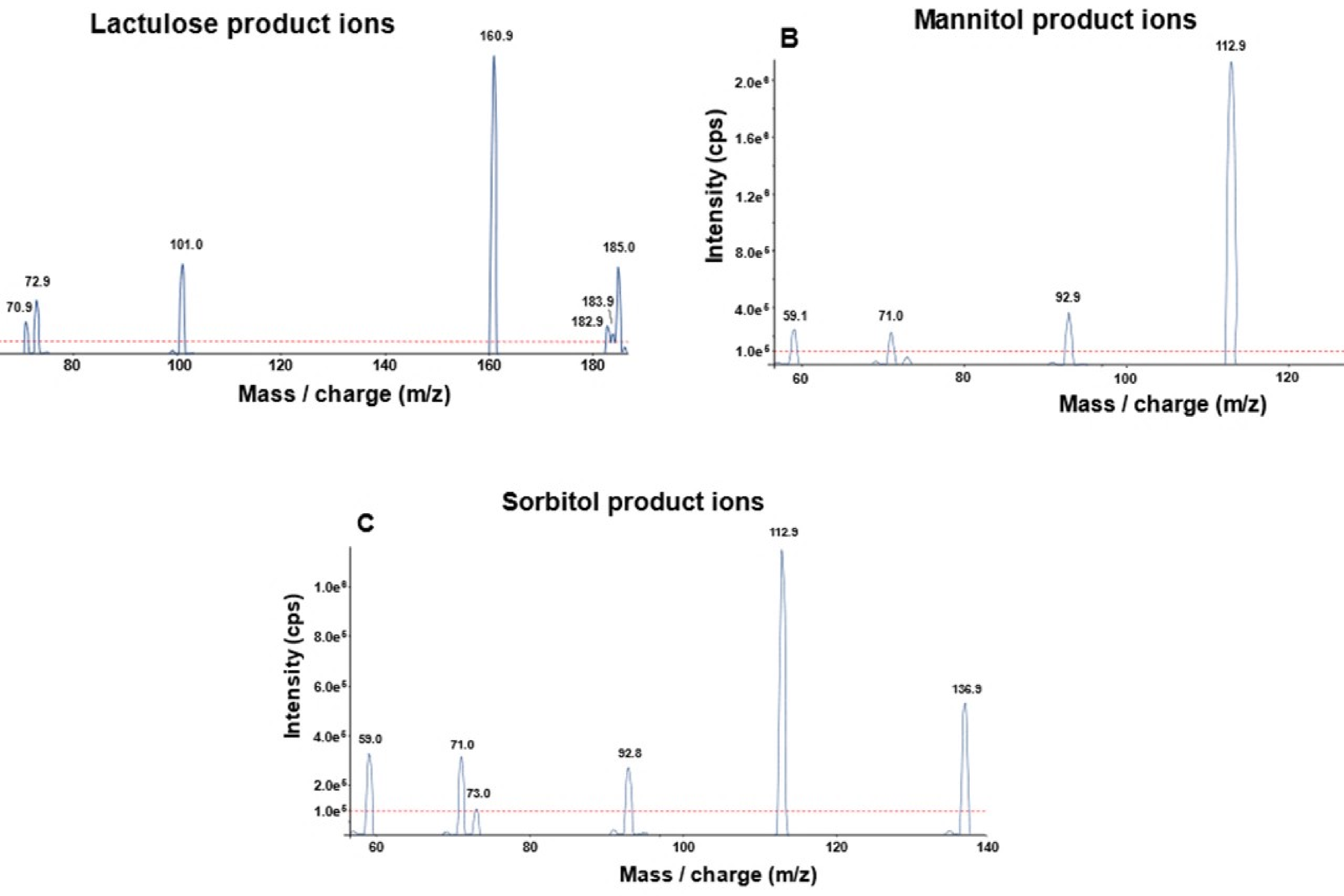
