## Supplementary material for "Measuring lactulose and mannitol levels using liquid chromatography coupled with tandem mass spectrum: application to clinical study of intestinal epithelium barrier function": Table, Supplement Digital Content A

**Table, Supplement Digital Content 1-** Mobile phase gradient for liquid chromatography system.

| Time (minutes) | Eluent A (%) | Eluent B (%) |
|---|---|---|
| 0 | 25 | 75 |
| 5 | 90 | 10 |
| 7 | 90 | 10 |
| 7.5 | 25 | 75 |
| 10 | 25 | 75 |

The run started with 75% acetonitrile in 0.05% formic acid (eluent B) at 25% $H_2O$ in 5mM ammonium acetate (eluent A; pH = 6.84) in 5 min, equilibrated for 2 min, returning to 75% eluent B in 0.5 min, and re-equilibrating for 2.5 min before the next injection.

**Table, Supplement Digital Content 4** - Mass spectrum monitoring and operational parameters of precursor ions and product ions of lactulose, mannitol, and sorbitol, through Multiple Reaction Monitoring (MRM).

| Analytes | Precursor ions (m/z) | Product ions (m/z) | DP [a] (V) [f] | EP [b] (V) | CE [c] (V) | CXP [d] (V) | Intensity [e] (cps) |
|---|---|---|---|---|---|---|---|
| **Lactulose** | 341.016 | 160.952<br>184.939<br>58.947<br>100.89<br>73.008 | -75 | -10 | -12<br>-48<br>-48<br>-22<br>-36 | -11<br>-3<br>-9<br>-9<br>-9 | 499000<br>420000<br>324000<br>301000<br>281000 |
| **Mannitol** | 180.932 | 112.798<br>136.813<br>58.994<br>71.009<br>89.033 | -60 | -10 | -10<br>-16<br>-26<br>-26<br>-18 | -9<br>-9<br>-9<br>-5<br>-7 | 230000<br>193000<br>124000<br>122000<br>109000 |
| **Sorbitol** | 180.935 | 112.912<br>58.924<br>136.927<br>70.973<br>92.879 | -75 | -10 | -26<br>-16<br>-26<br>-26<br>-24 | -9<br>-7<br>-9<br>-9<br>-11 | 225000<br>150000<br>143000<br>131000<br>107000 |

[a] Decomposition potential (voltage applied to the orifice to avoid clustering of ions); [b] entry potential; and [c] collision energy; [d] collision cell output potential; [e] intensity of fragment ions by MRM; [f] Volt (voltage measurement).

**Table, Supplement Digital Content 5-** Automatic parameters for precursor and product ions from lactulose, mannitol, and sorbitol, through flow injection analysis (FIA).

| Analytes | Precursor ions [a] (m/z) | Product ions [b] (m/z) | TEM [c] (ºC) [i] | $GS_1$ [d] (psi) [j] | CUR [e] (psi) | $GS_2$ [f] (psi) | IS [g] (V) [k] | CAD [h] |
|---|---|---|---|---|---|---|---|---|
| Lactulose | 341.016 | 160.952 | 500 | 50 | 20 | 40 | -3500 | Median |
| | | 184.939 | 700 | | 25 | 50 | | High |
| | | 58.947 | 450 | | 20 | 45 | | High |
| | | 100.89 | 550 | | 20 | 45 | | High |
| | | 73.008 | 450 | | 20 | 45 | | High |
| Mannitol | 180.932 | 112.798 | 450 | 50 | 20 | 50 | -4000 | Median |
| | | 136.813 | 700 | 45 | 30 | 45 | -4500 | Median |
| | | 58.994 | 500 | 50 | 22 | 50 | -4500 | Median |
| | | 71.009 | 550 | 50 | 22 | 50 | -4000 | High |
| | | 89.033 | 450 | 50 | 25 | 50 | -3500 | High |
| Sorbitol | 180.935 | 112.912 | 650 | 50 | 30 | 45 | -4500 | Median |
| | | 58.924 | 450 | 40 | 20 | 40 | -3500 | High |
| | | 136.927 | 650 | 50 | 20 | 50 | -4500 | High |
| | | 70.973 | 450 | 40 | 20 | 40 | -3500 | High |
| | | 92.879 | 700 | 40 | 20 | 45 | -4500 | Median |

[a] The Q precursor ion of analytes; [b] fragment ions; [c] temperature at the source; [d] nebulizer gas (helps in droplet formation); [e] gas flow between orifice and gas curtain; [f] heating gas (GS2 and TEM help with desolvation); [g] ions spray (voltage), which directs the formed ions to the analyzer; [h] gas collision (helps in the fragmentation of precursor ions to form product ions); [i] Celsius temperature unit; [j] pound-force per square inch; [k] voltage.

**Table 5** - Linearity, limit of detection (LD), limit of quantification (LQ) of the method in the LC-MS/MS system for analysis of the excretion of lactulose, mannitol and sorbitol sugars.

| **Analytes (precursor ion / product ions; m/z unit)** | **Calibration curve equation** | **LD [a] (ng/mL)** | **LQ [b] (ng/mL)** | ***R* [c]** |
|---|---|---|---|---|
| **Lactulose** (341.016 / 58.947) | $y = 1.15e^{4}x + 1.33e^{6}$ | 0.0055 | 0.0168 | 0.991 |
| **Mannitol** (180.932 / 71.009) | $y = 4.46e^{4}x + 6.42e^{6}$ | 0.0003 | 0.0010 | 0.993 |
| **Sorbitol** (180.935 / 58.924) | $y = 5.03e^{4}x + 7.10e^{6}$ | 0.0031 | 0.0001 | 0.995 |

[a] The detection limit; [b] limit of quantification; [c] correlation coefficient.

**Table, Supplement Digital Content 7**- Repeatability of the method in the LC-MS/MS system to analyze the excretion of lactulose, mannitol, and sorbitol sugars.

| | **Initial concentration [a] (ng/mL)** | **Concentration obtained [b] (ng/mL) (n = 3)** | **Recovery [c] (%)** | **SD [d]** | **CV [e] (%)** |
|---|---|---|---|---|---|
| **Lactulose** (341.016 / 58.947) | 100<br>500<br>1000 | 95.7<br>499.2<br>994.8 | 95.7<br>99.8<br>99.4 | 1.71<br>6.15<br>20.83 | 1.8<br>1.2<br>2.1 |
| **Mannitol** (180.932 / 71.009) | 100<br>500<br>1000 | 112.9<br>661.2<br>1116.4 | 112.9<br>132.2<br>111.6 | 11.06<br>8.63<br>18.71 | 9.8<br>1.3<br>1.7 |
| **Sorbitol** (180.935 / 58.924) | 100<br>500<br>1000 | 90.4<br>568.6<br>988.2 | 90.4<br>113.7<br>98.8 | 2.25<br>33.08<br>45.82 | 2.5<br>5.8<br>4.6 |

[a] Concentration in fortified samples; [b] concentration obtained through the average of the values obtained by the equation of the calibration curve of the spiked samples adding the standards in the samples of urine of volunteers; [c] % recovery; [d] standard deviation; and [e] coefficient of variation.

**Table, Supplement Digital Content 8-** Intermediate precision of the method in the LC-MS/MS system for analysis of lactulose, mannitol, and sorbitol in urine samples.

| Analytes (mass/charge; m/z) | Day | Initial concentration [a] (ng/mL) | Concentration obtained [a] (ng/mL) (n = 6) | Recovery [b] (%) | SD [c] | CV [d] (%) |
|---|---|---|---|---|---|---|
| **Lactulose** (341.016 / 58.947) | 1 | 500 | 621.6 | 124.3 | 41.1 | 6.6 |
| | 2 | | 504.7 | 100.4 | 11.8 | 2.3 |
| | 3 | | 607.7 | 121.5 | 52.3 | 8.6 |
| **Mannitol** (180.932 / 71.009) | 1 | 500 | 531.4 | 106.3 | 46.5 | 8.7 |
| | 2 | | 650.7 | 130.1 | 31.3 | 4.8 |
| | 3 | | 698.8 | 139.8 | 30.2 | 4.2 |
| **Sorbitol** (180.935 / 58.924) | 1 | 500 | 575.6 | 115.1 | 50.2 | 8.7 |
| | 2 | | 564.2 | 112.8 | 34.8 | 6.1 |
| | 3 | | 657.8 | 131.6 | 104.0 | 5.8 |

[a] Concentration in fortified samples; [b] percentage of recovery of concentrations obtained through the equation of the calibration curve in fortified samples adding the standards in the samples of urine of volunteers; [c] % recovery of mean concentrations obtained on different days; [d] standard deviation; and coefficient of variation.

**Table, Supplement Digital Content 9**- Intensity of lactulose and mannitol analytes in standard solutions and fortified urine samples. Fortified urine samples (FU) obtained from known concentrations of analytes in urine samples.

| Analytes | Concentration [a] (ng/mL) | Samples | Intensity of analytes [b] (cps; N = 3) | SD [c] | CV [d] (%) |
|---|---|---|---|---|---|
| Lactulose | 750 | FU | 54176.67 | 1552.23 | 3.0 |
| | | Standard | 51866.67 | 1521.26 | 2.9 |
| | 1500 | UF | 76046.67 | 5877.77 | 6.6 |
| | | Standard | 95003.33 | 8227.71 | 8.7 |
| | 3000 | FU | 130410.00 | 10424.17 | 6.7 |
| | | Standard | 174466.67 | 21185.92 | 12.1 |
| | 6000 | UF | 245300.00 | 7031.59 | 2.5 |
| | | Standard | 383666.67 | 31187.55 | 8.1 |
| Mannitol | 750 | FU | 12523333.33 | 519839.71 | 4.2 |
| | | Standard | 23706666.67 | 973721.38 | 6.6 |
| | 1500 | FU | 13000000.00 | 253245.59 | 1.1 |
| | | Standard | 24000000.00 | 1769755.16 | 6.3 |
| | 3000 | FU | 41350000.00 | 727461.33 | 1.8 |
| | | Standard | 49000000.00 | 1545930.14 | 3.2 |
| | 6000 | FU | 71253333.33 | 1912598.58 | 2.7 |
| | | Standard | 84000000.00 | 1553093.68 | 1.8 |

[a] Concentration of analytes in standard solutions and in fortified urine; [b] mean intensities of lactulose and mannitol analytes; [c] standard deviation; [d] coefficient of variation.

**Table, Supplement Digital Content 10-** Retention time of lactulose and mannitol in standard solutions and fortified urine samples. Retention time reproduced on standard and fortified urine (FU) samples.

| Analyte's concentrations (ng/mL) | Analytes | | | | | |
|---|---|---|---|---|---|---|
| | Mannitol | | | Lactulose | | |
| | Retention time [a] (min) | Retention time FU [b] (min) | Recovery (%) [c] | Retention time (min) | Retention time FU (min) | Recovery (%) |
| 750 | 6.87 | 6.83 | 99.4 | 7.18 | 7.13 | 99.3 |
| 1500 | 6.86 | 6.83 | 99.6 | 7.17 | 7.14 | 99.6 |
| 3000 | 6.84 | 6.82 | 99.7 | 7.16 | 7.13 | 99.6 |
| 6000 | 6.84 | 6.82 | 99.7 | 7.16 | 7.12 | 99.4 |
| 750 | 6.81 | 6.81 | 100.0 | 7.13 | 7.12 | 99.9 |
| 1500 | 6.82 | 6.81 | 99.9 | 7.14 | 7.12 | 99.7 |
| 3000 | 6.81 | 6.81 | 100.0 | 7.13 | 7.12 | 99.9 |
| 6000 | 6.81 | 6.80 | 99.9 | 7.14 | 7.11 | 99.6 |
| 750 | 6.8 | 6.84 | 100.6 | 7.12 | 7.14 | 100.3 |
| 1500 | 6.81 | 6.81 | 100.0 | 7.12 | 7.12 | 100.0 |
| 3000 | 6.82 | 6.81 | 99.9 | 7.14 | 7.12 | 99.7 |
| 6000 | 6.81 | 6.81 | 100.0 | 7.13 | 7.11 | 99.7 |

[a] Retention time obtained with the standard solution; [b] retention time obtained with the fortified urine; [c] % recovery.

**Table, Supplement Digital Content 11** - Recovery and matrix effect of lactulose and mannitol in fortified urine. Fortified urine obtained by adding known concentrations of the analyte to the urine of volunteers in triplicate.

| Analytes | Concentration [a] (ng/mL) | Recovery [b] (%) | EM [c] (%) |
|---|---|---|---|
| **Lactulose** | 750 | 99.8 | 4.4 |
| | 1500 | 93.9 | -8.9 |
| | 3000 | 88.8 | -22.0 |
| | 6000 | 135.3 | -30.8 |
| **Mannitol** | 750 | 84.7 | -11.7 |
| | 1500 | 84.2 | -10.8 |
| | 3000 | 85.2 | -11.8 |
| | 6000 | 84.4 | -12.4 |

[a] Known Concentration added to standard solutions and urine samples; [b] percentage obtained through the average of analyte concentrations; [c] percentage expressed in suppression of analytes concentrations.

**Table, Supplement Digital Content 13** – Unidirectional analysis of covariance (ANCOVA) between the experimental groups, independent variable, and in the dependent variable, lactulose:mannitol excretion rate (Napierian logarithm of lactulose:mannitol rate) to correct the covariate age of children.

| Source | Type III Sum of Squares | df | Mean Square | F | Sig. | Partial Eta Squared |
|---|---|---|---|---|---|---|
| **Corrected Model** | 7.569 [a] | 3 | 2.523 | 11.812 | 0.000 | 0.330 |
| **Intercept** | 83.187 | 1 | 83.187 | 389.479 | 0.000 | 0.844 |
| **Age (months)** | 0.000 | 1 | 0.000 | 0.002 | 0.966 | 0.000 |
| **Groups** | 3.134 | 2 | 1.567 | 7.336 | 0.001 | 0.169 |
| **Error** | 15.378 | 72 | 0.214 | | | |
| **Total** | 365.935 | 76 | | | | |
| **Corrected Total** | 22.947 | 75 | | | | |

[a] R Squared = 0.330 (Adjusted R Squared = 0.302).

**Text, Supplement Digital Content 14**

The mass analyzer used in this work was a triple quadrupole, being the most used in quantitative analysis (1). The ionization method used determines how the sample material is transferred to the mass spectrometer (2). The ionization source was electrospray ionization (ESI), being the most used source for coupling with the mass spectrum (2,3). Another advantage of the HLPC-MS/MS system is the speed in the individual analysis of the samples when compared to the liquid chromatography system with pulsed amperometric detection (HPLC-PAD), where the run lasts 35 minutes and in the present system, only 10 minutes. Thus, reducing the race time to less than a third. With this, a greater number of runs can be done in a shorter time interval, thus increasing the dosage robustness in the LC-MS/MS platform, giving another advantage to this system.

Due to the characteristics of the described process, ESI applies very well to compounds of medium and high polarity, hence its extensive applicability in the pharmaceutical, food, natural products, among others (4-6). The automatic acquisition mode used was the Multiple Reaction Monitoring (MRM), indicated for quantitative analysis (7). The first step in developing the MRM method is to determine the best conditions for analyzing the analytes of interest, as described by Iglesias (8). The ability of MS/MS to monitor multiple reaction ion transitions per single run gives the method high specificity (9). We observed the MS/MS method to be highly specific for the detection of single product ions for lactulose, mannitol, and sorbitol, as demonstrated in this work and reported by other authors (9,10). The five selected product ions were obtained from high collision energies. According to Kind et. al. (2), low collision energies preserve precursor ions and only a few product ions are observed. Increasing the collision energy promotes the abundance of product ions towards lower m/z ranges and at the same time decreases the abundance of the precursor ion.

The HILIC-ZIC® column was chosen because it offers greater retention of compounds such as lactulose, mannitol, and sorbitol, as described by several authors (9,11). The HILIC-ZIC® column also offered satisfactory separation and reproducible retention times for lactulose, mannitol and sorbitol in the standards and analyzed samples as reported by Kubica et al. (9). Liquid chromatography with the HILIC-ZIC® column can separate polar compounds with the same molar mass and mass spectrometry is able to identify them (9,10). We were able to observe the separation of mannitol and sorbitol compounds, even with the same molar masses, due to the ability of mass spectrometry to separate through the mass/charge ratio (m/z) and the precursor ion / product ion ratio in the triple quadrupole, confirming the advantage of the LC-MS/MS system against the isolated HPLC-PAD in the specification of the analyte of interest. The column provided reproducible separation and retention time, as reported by Kubica et al. (9). The use of retention time is highly recommended for high confidence compound identification (12,13).

We observed better ionization of ions through the negative mode using formic acid as an additive compound. Formic acid is known as a positive mode additive, but it can also be used as a negative ionization mode additive (14). 0.1% formic acid is considered a mobile phase additive aiding the ionization of analytes. It is volatile and must be used in low concentration, to avoid interference in the analyte ionization process, a process known as ionization suppression, as described by other authors such as Iglesias (8) and Trufelli et al. (15).

The presence of formic acid (FA) in the mobile phase under different concentrations changed its pH, varying according to the added concentration. The higher the concentration of AF added, the lower the pH of the mobile phase. Mobile phase with pH below 6 did not show good ionization. The mobile phase with the addition of 0.05% of FA showed better ionization and pH close to neutral (pH = 6.84). Gan et al. (16) also observed that changes in pH or organic percentage can significantly increase ESI ionization. According to Van Wijck et al. (17), the application of a weaker acid, such as formic acid, increases the disaccharide response, but strongly reduces the monosaccharide response. Differently from the above, we found in the presence of AF 0.05% an improvement in the response of both lactulose and mannitol.

During the mobile phase definition process, we identified several changes in ionization according to the mobile phase gradient. There were several attempts until we arrived at the gradient that would provide the best ionization. As electrospray efficiency depends primarily on mobile phase composition, the optimal eluent composition for proper chromatographic separation is sometimes inadequate to achieve maximum electrospray response (18). As described by Lostia et al. (10), the present method, based on fragment ion identification, overcomes many of the problems encountered in the analysis of carbohydrates in biological fluids and can be considered a useful automated tool to study intestinal functions and the modification of the functional gastrointestinal barrier, both in pediatrics and also in adult diseases such as celiac disease, Crohn's disease, and inflammatory bowel disease, allowing accurate patient discrimination also for dietary restrictions.

The matrix effect should always be evaluated when validating quantitative LC-MS/MS methods, especially in complex matrices such as urine (19). One of the biggest problems of analysis through mass spectrometry is to minimize the matrix effect (20). One of the strategies adopted that can minimize this effect is sample purification or a more efficient chromatographic separation (21). We performed the matrix effect interference analysis by the reference chemical substance (SQR) addition method, in which known amounts of SQR were added to the sample. We observed that the retention time of the analytes remained without significant variations, within the range of 2% of the times in the standard as described by Núñez, Moyano and Galceran (18). The recovery ranged between 84.23 and 135.27.

We observed the suppression of ionization at almost all concentrations, being greater at the concentration of 6000 ng/mL in the three analytes. In our method the reported ion suppression remained between 8-30%. However, suppression did not affect the accuracy and precision of the assay as the signals for lactulose and mannitol were above the signals for LD and LQ (19).

---

**Figure legend, Supplement Digital Content 2**

Identification of lactulose, mannitol, and sorbitol precursor ions through automatic optimization using the Multiple Reaction Monitoring (MRM) method in the QTRAP® 5500. Lactulose, mannitol, and sorbitol standards were entered directly into the mass spectrometer. As expected, in the sample without the addition of standard sugars, precursor ions (A) were not identified. Using the MRM method, it was possible to identify the precursor ions of lactulose (B), mannitol (C) and sorbitol (D).

**Figure legend, Supplement Digital Content 3**

**Spectrograms obtained in negative ionization mode for lactulose, mannitol, and sorbitol product ions**. Product ion intensity data were obtained by direct injection into the QTRAP 5500 mass spectrometer (ABSciex (Framingham, MA, USA) at a concentration of 10 ng/mL The spectrograms show the five main product ions of each of the lactulose (**A**), mannitol (**B**), and sorbitol (**C**), respectively, of higher intensity selected in the third pole of the quadrupole system of the mass spectrometer. The red dashed line indicates the minimum intensity limit (cps) acceptable for the product ions.

**Figure legend, Supplement Digital Content 12**

**Map of Fortaleza city, state of Ceará, Brazil and study location**. This figure shows the geographic location of the city of Fortaleza on the map of Brazil and the heat map of Fortaleza with the human development index (HDI). The dots marked in the map represent the origin of children recruited in the case-control study, from August 19, 2010, to May 23, 2017.
