## Supplementary material for "Measuring lactulose and mannitol levels using liquid chromatography coupled with tandem mass spectrum: application to clinical study of intestinal epithelium barrier function": Table, Supplement Digital Content B

**Table B** Automatic parameters for precursor and product ions from lactulose, mannitol, and sorbitol, through flow injection analysis (FIA).

| Analytes | Precursor ions [a] (m/z) | Product ions [b] (m/z) | TEM [c] (ºC) [i] | $GS_1$ [d] (psi) [j] | CUR [e] (psi) | $GS_2$ [f] (psi) | IS [g] (V) [k] | CAD [h] |
|---|---|---|---|---|---|---|---|---|
| | | 160.952 | 500 | | 20 | 40 | | Median |
| | | 184.939 | 700 | | 25 | 50 | | High |
| Lactulose | 341.016 | 58.947 | 450 | 50 | 20 | 45 | -3500 | High |
| | | 100.89 | 550 | | 20 | 45 | | High |
| | | 73.008 | 450 | | 20 | 45 | | High |
| | | 112.798 | 450 | 50 | 20 | 50 | -4000 | Median |
| | | 136.813 | 700 | 45 | 30 | 45 | -4500 | Median |
| Mannitol | 180.932 | 58.994 | 500 | 50 | 22 | 50 | -4500 | Median |
| | | 71.009 | 550 | 50 | 22 | 50 | -4000 | High |
| | | 89.033 | 450 | 50 | 25 | 50 | -3500 | High |
| | | 112.912 | 650 | 50 | 30 | 45 | -4500 | Median |
| | | 58.924 | 450 | 40 | 20 | 40 | -3500 | High |
| Sorbitol | 180.935 | 136.927 | 650 | 50 | 20 | 50 | -4500 | High |
| | | 70.973 | 450 | 40 | 20 | 40 | -3500 | High |
| | | 92.879 | 700 | 40 | 20 | 45 | -4500 | Median |

[a] The Q precursor ion of analytes; [b] fragment ions; [c] temperature at the source; [d] nebulizer gas (helps in droplet formation); [e] gas flow between orifice and gas curtain; [f] heating gas (GS2 and TEM help with desolvation); [g] ions spray (voltage), which directs the formed ions to the analyzer; [h] gas collision (helps in the fragmentation of precursor ions to form product ions); [i] Celsius temperature unit; [j] pound-force per square inch; [k] voltage.
