## Supplementary material for "Measuring lactulose and mannitol levels using liquid chromatography coupled with tandem mass spectrum: application to clinical study of intestinal epithelium barrier function": Table, Supplement Digital Content E

**Table, Supplement Digital Content E** – Unidirectional analysis of covariance (ANCOVA) between the experimental groups, independent variable, and in the dependent variable, lactulose:mannitol excretion rate (Napierian logarithm of lactulose:mannitol rate) to correct the covariate age of children.

| Source | Type III Sum of Squares | df | Mean Square | F | Sig. | Partial Eta Squared |
|---|---|---|---|---|---|---|
| **Corrected Model** | 5,824[a] | 2 | 2,912 | 18,365 | ,000 | ,542 |
| **Intercept** | 54,566 | 1 | 54,566 | 344,130 | ,000 | ,917 |
| **Agem** | ,002 | 1 | ,002 | ,015 | ,904 | ,000 |
| **CEFCC** | 3,166 | 1 | 3,166 | 19,967 | ,000 | ,392 |
| **Error** | 4,915 | 31 | ,159 | | | |
| **Total** | 203,211 | 34 | | | | |
| **Corrected Total** | 10,739 | 33 | | | | |

a. R Squared = ,542 (Adjusted R Squared = ,513)
